# Insight into vitreous biomechanics and clinical implications: A systematic review

**DOI:** 10.1101/2025.09.17.25335952

**Authors:** Lauren F Ong, Tara Suresh, Esther Y Son, Dan Schwartz, Frank Brodie

## Abstract

**Purpose:** This systematic review investigates the current landscape of rheometric techniques used to study the viscosity and viscoelastic properties of vitreous humor to better understand vitreous behavior for clinical applications, particularly in anomalous posterior vitreous detachment (PVD).

**Methods:** A comprehensive literature search was conducted in PubMed, EMBASE, and Scopus using the keywords “vitreous body,” “viscosity,” and “rheology.” The review followed PRISMA guidelines, including studies assessing vitreous viscosity in human or animal eyes with experimental methods. Non-peer-reviewed and non-English studies were excluded. The quality of the 42 studies selected was assessed using the SYRCLE and adapted Newcastle-Ottawa Scale (NOS) tools. Rheometric devices, experimental models, and outcomes were analyzed.

**Results:** The 42 studies analyzed 1,125 experimental eyes, including porcine (33.3%), bovine (25%), and human (15%) eyes. Various devices were used for experimentation with capillary tubes (20%) and rotational devices (15%) being most common. Mechanical properties such as G’ and G’’ were reported in 10 and 8 studies, respectively. Steady-state viscosity (ηss) was reported in 15 studies, values ranging from 1.43 × 10^5^ ± 2.19 × 10^4^ Pa·s to 12.3 ± 0.94 Pa·s across studies.

**Conclusions:** This systematic review highlights variability in rheometric methods with species, techniques, and outcomes to assess vitreous humor. Standardized methods are necessary to assess vitreous properties and for clinical application.

## Introduction

Vitreous humor, the gel-like substance that fills the human eye, plays a critical role in maintaining ocular structure and function [1]. As a dynamic, viscoelastic material, the vitreous undergoes natural liquefaction with aging, a process that affects its viscosity and elasticity [2]. These changes can contribute to the development of various retinal pathologies, including anomalous posterior vitreous detachment (PVD), vitreo-macular traction (VMT), macular holes, and retinal tears [3,4].

Despite its clinical significance, the process of vitreous liquefaction and its role in the pathogenesis of vitreous-related diseases remains poorly understood [5,6]. Rheometry has emerged as a powerful tool to study the viscoelastic properties of vitreous humor. However, significant heterogeneity exists in both the methods used and the experimental models studied [7]. This variability, as well as the scarcity of human studies, limits our understanding of the fundamental physiology of the vitreous.

A better understanding of vitreous viscosity has the potential to inform surgical planning in vitreous- related pathologies such as macular holes, retinal detachment, and PVD [2,8,9], optimize intravitreal drug delivery for conditions like diabetic macular edema, age-related macular degeneration, and retinal vein occlusions [10–12], and guide the development of vitreous substitutes that mimic the biomechanical properties of the natural vitreous [9,13,14]. This systematic review aims to provide a comprehensive synthesis of the existing literature on rheometric techniques used to assess vitreous viscosity, with a focus on the key experimental findings. Additionally, this review will address the gaps in knowledge related to vitreous viscosity, particularly in human eyes, and will propose future directions for research and technological development.

## Materials and Methods

### Search Strategy

This systematic review was conducted in adherence with the PRISMA (Preferred Reporting Items for Systematic Reviews and Meta-Analyses) guidelines and adhered to a protocol registered with PROSPERO (CRD42024495414). A comprehensive search was conducted using PubMed, EMBASE, and Scopus databases. Search terms were developed based on the Population, Intervention, Comparator, Outcomes, Timing, and Study Design (PICOTS) framework (**Table 1**). Combinations of keywords such as “vitreous humor,” “viscosity,” “rheometry,” and specific animal models were used with Boolean operators “AND” and “OR.” The detailed search strategy and PRISMA checklist are provided in **Appendix S1 and Appendix S2**, respectively.

**Table 1.** PICOTS Eligibility Criteria. The Population, Intervention, Comparator, Outcomes, Timing, and Study Design (PICOTS) framework used to define inclusion and exclusion criteria.

|  |  |
| --- | --- |
| **Population** | **Inclusion**<br>• Studies investigating the vitreous humor viscosity in animal models of any species.<br>**Exclusion**<br>• Studies that do not examine vitreous humor or focus solely on computational models. |
| **Intervention** | **Inclusion**<br>• Rheometric analysis of vitreous humor using techniques such as rotational viscometry or oscillatory rheometry.<br>**Exclusion**<br>• Studies not using rheometric techniques to evaluate vitreous humor. |
| **Comparator** | • Studies with or without comparators (e.g., comparisons between species or pathological conditions). |
| **Outcomes** | **Inclusion**<br>• Outcomes analyzed:<br>◦ Quantitative viscosity measurements<br>◦ Rheological properties<br>◦ Qualitative descriptions of vitreous behavior<br>• Studies not reporting viscosity-related outcomes were excluded. |
| **Timing** | • Any experimental timeframe or duration of viscosity analysis. |
| **Setting** | **Inclusion**<br>• Laboratory-based studies with animal models.<br>**Exclusion**<br>• In silico or computational-only studies; studies without laboratory-based rheometric analysis. |

### Inclusion and Exclusion Criteria

Inclusion criteria were as follows: 1) studies using experimental techniques to evaluate the viscosity of vitreous humor in animal eyes; 2) studies reporting quantitative or qualitative viscosity outcomes; and 3) peer-reviewed, full-length articles published in English.

Exclusion criteria were as follows: 1) studies not investigating the viscosity of vitreous humor; 2) in silico studies or studies without experimental data; 3) articles not available in full-text or not peer-reviewed; 4) studies not published in English.

### Study Selection

Covidence software was utilized to facilitate the screening and selection process. Two reviewers independently performed a dual, blinded review of titles and abstracts against the inclusion and exclusion criteria. Articles meeting these criteria advanced to full-text review. During the full-text screening phase, reasons for exclusion were documented. Any disagreements at any stage were resolved by group consensus with a “gold standard” reviewer (LO or FB). A PRISMA flow diagram was generated to document the study selection process.

### Data Extraction and Analysis

Forty-two full-text articles were selected for focused review. Data extraction was performed independently by two reviewers using a standardized abstraction form. The extracted data included:

● Study characteristics: author and year
● Animal or human models: species used in the experiments.
● Experimental techniques: rheometric device name, type (e.g., geometry), controlled variables (e.g., flow, shear rate).
● Primary outcomes: viscosity values, units of measurement, and experimental findings.

Given the heterogeneity of experimental methods and outcomes, a narrative synthesis was conducted. Key findings were compared across techniques and species to evaluate patterns and differences in the reported outcomes.

### Quality Assessment

The quality of included studies was assessed independently by two reviewers. Controlled studies with separate treatment arms were evaluated using SYRCLE’s (Systematic Review Centre for Laboratory Animal Experimentation) risk of bias tool, which examines sequence generation, allocation concealment, blinding, and other domains [15]. For non-controlled study designs such as cohort and case-control studies, the Newcastle-Ottawa Scale (NOS) was adapted for animal studies to assess study quality based on criteria such as selection, comparability, and outcomes [16]. Remaining study designs that did not fall under the above risk of bias tools were discussed narratively. Disagreements were resolved through consensus. A detailed assessment of the risk of bias for all studies is provided in **Appendix S3**.

## Results

### Literature Selection

As outlined in **Figure 1**, our initial search using PubMed, Scopus, and Embase yielded 2,401 articles to screen. After removing 459 duplicates, 1,942 articles remained for further evaluation. During the title and abstract screening phase, 1,822 articles were excluded. A total of 78 articles were removed during the full-text screening. Of these, 27 were excluded because they did not report vitreous humor viscosity outcomes, 17 were removed for being non-English articles, 15 were removed for lacking peer-reviewed full-length articles, 14 were removed for lacking test subjects undergoing experimental techniques, and 5 were excluded for not being original studies.

**Figure 1.**
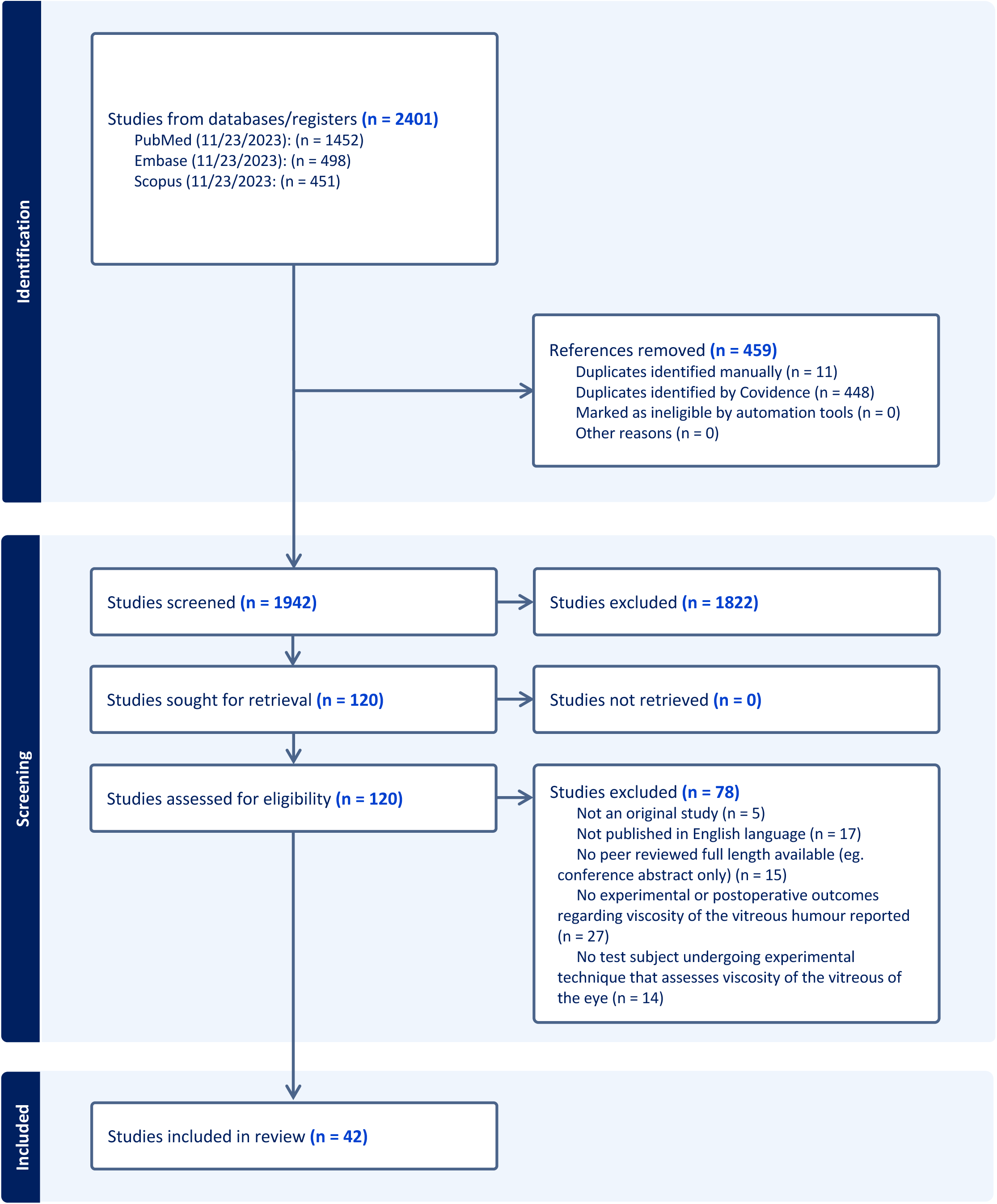
PRISMA Flow Diagram. A flowchart illustrating the systematic review process, including the number of studies identified, screened, excluded, and included at each stage.

From the initial 2,401 articles, 42 studies met the inclusion criteria for this systematic review [17–58]. Of the papers that reported numeric counts, a total of 1,125 eyes were analyzed in these studies. Twenty-two papers did not report counts of eyes used for their experiments, and 2 papers reported partial counts for eyes. The primary species studied included bovine, porcine, human, ovine, and other miscellaneous groups. The viscosity measurements reported in these studies included both steady-state and dynamic viscosities, with some studies using additional methods such as complex viscosity. A detailed summary of all the included studies is shown in **Appendix S4**.

### Publication Year

Studies were published between the years 1958 and 2022, with the largest number of studies published prior to 2000 (n = 15, 35.7%) and between 2011–2015 (n = 13, 31.0%). Fewer studies were published in the periods of 2001–2005 (n = 1, 2.4%), 2006–2010 (n = 4, 9.5%), 2016–2020 (n = 7, 16.7%), and 2021–2023 (n = 2, 4.8%) (**Figure 2**).

**Figure 2.**
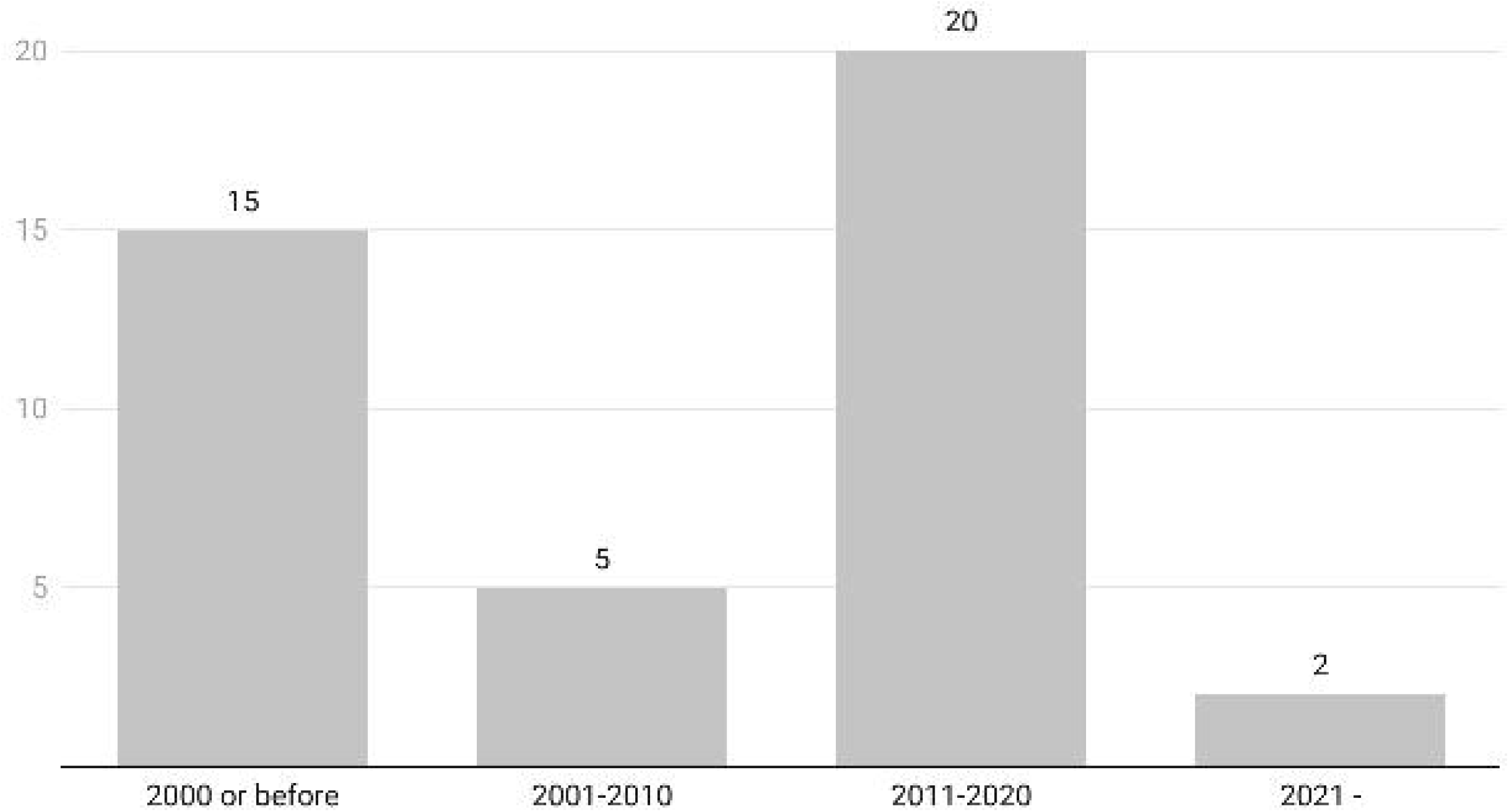
Publication Years of Included Papers. Distribution of included studies by publication year, highlighting trends over time.

### Device Types

A variety of devices were employed in the studies, with the most common being capillary tubes (n = 9, 20%) and rotational devices (n = 7, 15%). Other frequently used devices included cone-plate systems (n = 4, 9%) and parallel plate systems (n = 4, 9%). Several unique devices, such as torsion pendulum apparatus, viscoelastometers, and colloidal cantilevers, were each reported in a single study (2%). A comprehensive list of devices and their frequencies is shown in **Table 2**.

**Table 2.** Study Rheometrical Devices. A summary of the different rheometric devices utilized in the included studies, listing their frequency and types (e.g., capillary tubes, rotational devices, cone-plate systems).

| Device Types (n=24) | Experiments n (%) ▼ |
| --- | --- |
| Capillary tube | 9 (20) |
| Rotational | 7 (15) |
| Cone-plate | 4 (9) |
| Parallel plate | 4 (9) |
| Probe | 2 (4) |
| Rotational and cone-plate attachment | 2 (4) |
| Cleat geometry | 1 (2) |
| Colloidal cantilever | 1 (2) |
| Computer analysis of the resonant curves | 1 (2) |
| Concentric cylinder | 1 (2) |
| Cylindrical probe | 1 (2) |
| Fluorescence Recovery After Photobleaching | 1 (2) |
| Horizontal viscosimeter | 1 (2) |
| Laser speckle rheology | 1 (2) |
| Magnet | 1 (2) |
| Magnetic microprobe | 1 (2) |
| Magnetic robot | 1 (2) |
| Magnetic resonance imaging | 1 (2) |
| Microbubble | 1 (2) |
| Modified viscometer | 1 (2) |
| Oscillation | 1 (2) |
| Rotational with parallel plate geometry | 1 (2) |
| Torsion pendulum apparatus | 1 (2) |
| Viscoelastometer | 1 (2) |

### Experimental Models

The most frequently used experimental models were porcine (n = 20, 33.3%)[27,29,30,33,35,37,39,41,43–48,50,51,54,55,57] and bovine/calf/cattle eyes (n = 15, 25.0%)[19,21,23,27,29,31,32,37,39,45,46,53,58]. Human eyes were used in 9 studies (15.0%)[22,33,35,36,40,42,45,46], followed by rabbit eyes (n = 8, 13.3%)[17,18,24,28,34,37,42,52]. Less common models included goat (n = 2, 3.3%)[37,49], monkey (n = 2, 3.3%)[25,52], sheep (n = 2, 3.3%)[26,37,46], owl (n = 1, 1.7%)[20], and ox (n = 1, 1.7%)[56]. The distribution of experimental models is illustrated in **Figure 3**.

**Figure 3.**
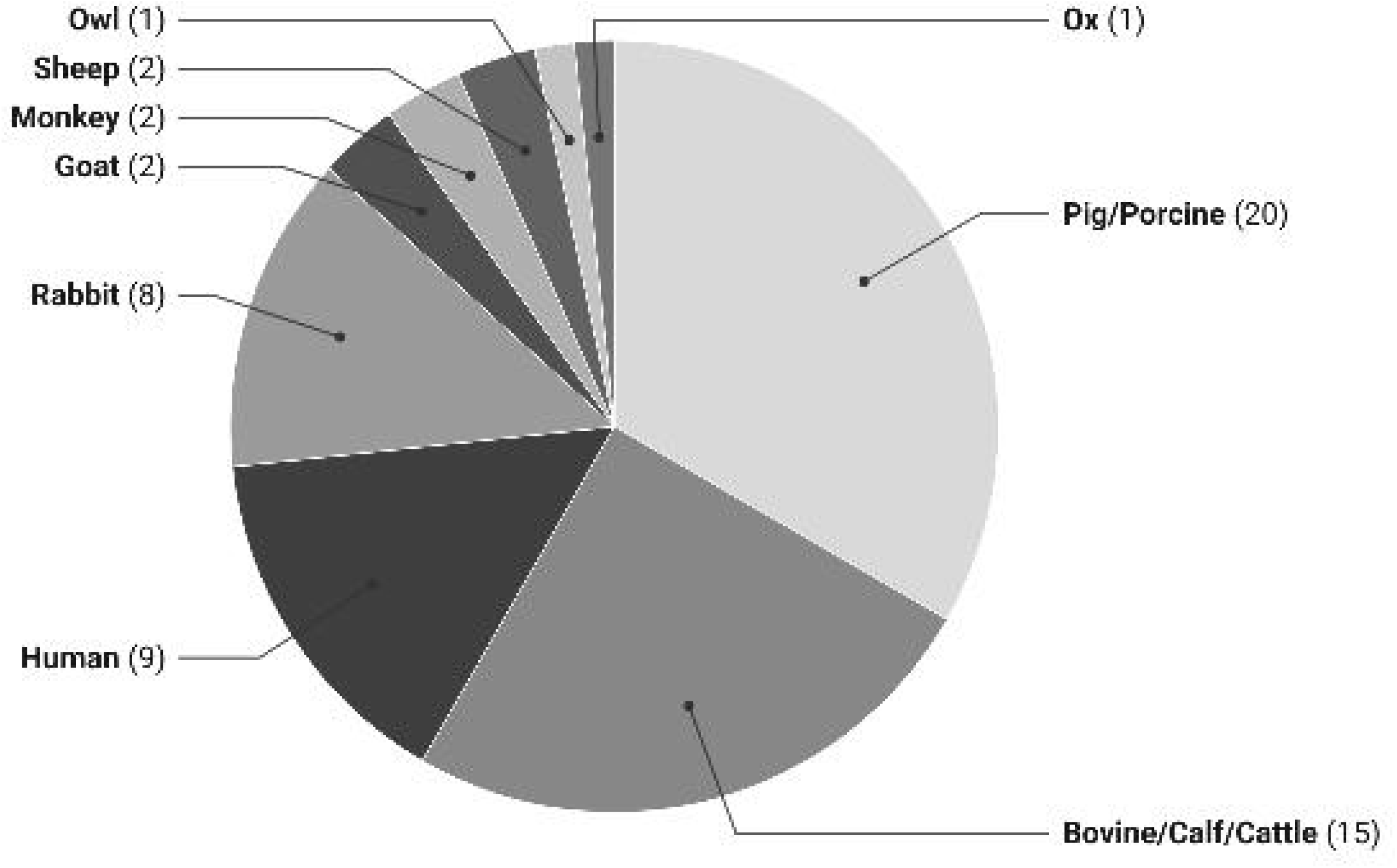
Experimental models. Distribution of animal and human models used in the included studies, reported as counts.

### Human Eyes

Nine studies measured the viscosity of human vitreous humor using various methods and devices (**Table 3**)[21,33,35,36,38,40,42,45,46]. In 1964, Berman and Michaelson measured the relative viscosity of post-mortem human eyes using a Cannon-Manning semimicro capillary viscometer, reporting a viscosity of 1.04 for infants (0-2 years), 1.53 for individuals aged 13-45 years, and 2.08 for those aged 50-85 years, with myopic eyes showing a viscosity of 1.54 [21]. Locke and Morton (1965) analyzed 176 human eye samples with an Ostwald-type viscosimeter, finding an average viscosity of 1.97, with a range from 10.6 to 5.50 [36]. Kawano et al.employed a Cannon-Fenske kinematic viscometer and cone-plate rotary viscometer in 1982, confirming that the addition of 0.005% thimerosal did not affect the viscosity of the vitreous [33].

**Table 3.**
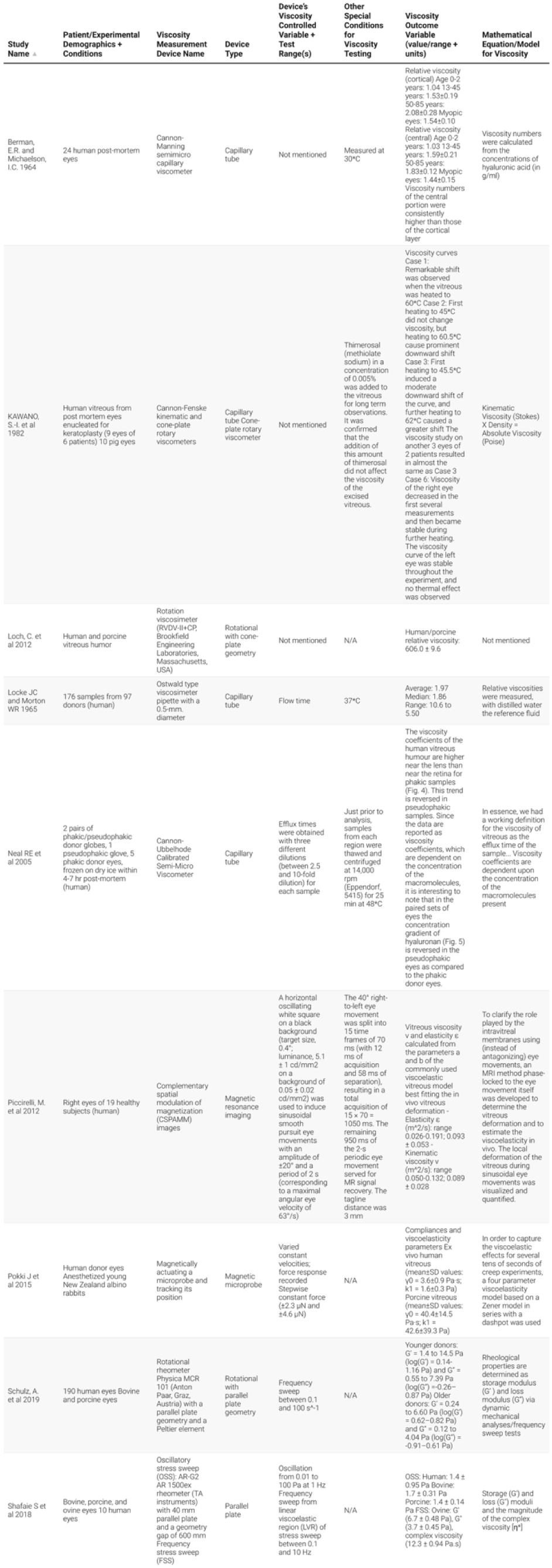
Viscosity Measurements in Human Eyes. A table summarizing studies that measured the viscosity of human vitreous humor, including details on measurement techniques, devices used, and key findings.

Over 30 years later in 2005, Neal et al. used a Cannon-Ubbelhode calibrated semi-micro viscometer to measure efflux times in human vitreous from 5 phakic donor eyes, noting that viscosity coefficients depended on the concentration of macromolecules present in the samples [38]. Pokki et al. employed a magnetic microprobe for human vitreous, reporting a viscosity of 3.6±0.9 Pa·s [42]. Shafaie et al. used an oscillatory stress sweep test with a parallel plate and found a viscosity of 1.4±0.95 Pa for human vitreous [46]. Piccirelli et al. utilized complementary spatial modulation of magnetization (CSPAMM) imaging to assess the viscosity of the vitreous humor in the right eyes of 19 healthy human subjects [40]. Most recently in 2019, Schulz et al. measured the rheological properties of human vitreous using a rotational rheometer with parallel plate geometry, reporting storage modulus (G’) values ranging from 0.14 to 1.16 Pa in younger donors and 0.62 to 0.82 Pa in older donors [45]

### Vitreous Humor Mechanical Properties

#### Storage Modulus (G’) and Loss Modulus (G’’)

The storage modulus (G’) and loss modulus (G’’) were reported in pascals (Pa). G’ values were reported in 10 studies, ranging from 0.24 Pa to 14.5 Pa [32,37,39,45–47,50,51,57,58]. G’’ values were reported in Pa in 8 studies, ranging from 0.12 Pa to 7.39 Pa [37,39,45–47,50,51,57].

#### Viscosity (ηss)

Steady-state viscosity (ηss) was reported in pascal-seconds (Pa·s) or as relative viscosities in 15 studies [17–22,25,27,34–36,42–44,56]. Examples of viscosity values reported in Pa·s include 1.43 × 10^5 ± 2.19 × 10^4 Pa·s (Connelly et al., 2016), 4.35 × 10^4 ± 4.80 × 10^3 Pa·s (Connelly et al., 2016), and 12.3 ± 0.94 Pa·s (FSS, 2017).

#### Other Units of Measurement

Other units of measurement used in two studies include centipoise (cp) and poise (P). Viscosity was reported in centipoise (cP) in 3 studies [28,30,49]. Gisladottir et al., 2009 (6.29 ± 2.3 cp) and Loch et al., 2012 (606.0 ± 9.6 cp). The unit poise (P), where 1 P = 0.1 Pa·s, was used in some studies for viscosity measurements.

### Risk of Bias

Risk of bias, assessed using the adapted Newcastle-Ottawa Scale (NOS) for animal studies as no studies had separate treatment arms warranting the use of SYRCLE’s risk of bias tool, was generally moderate across the included studies. The domain most frequently rated as “unclear” was whether alternative explanations for the observed effects were ruled out, such as through control groups, randomization, or blinding. Reporting of methodological detail was mixed: some studies provided complete descriptions of species, strain, sex, housing conditions, and intervention protocols, while others gave insufficient information to ensure reproducibility in other research settings. Other NOS domains were generally judged as clear of bias. Full NOS scoring for each study is presented in **Appendix S3**.

## Discussion

### Gaps in Knowledge

The results of this systematic review underscore the substantial gaps in our understanding of vitreous viscosity, particularly in human eyes. Although rheometric techniques have been used extensively in animal models, the majority of the studies in this review involved porcine (33%) and bovine (25%) eyes, with human eyes comprising only 15% of the total experimental studies. The differences in vitreous physiology between these species and humans could introduce significant variability in the findings, limiting the applicability of animal model data to human clinical settings.

Animal eyes, while offering useful insights into the basic mechanics of vitreous behaviour, may not fully replicate the viscoelastic properties or age-related liquefaction processes seen in human vitreous. Human vitreous humor is distinct in its composition, containing higher concentrations of hyaluronic acid and collagen fibers, which may affect its viscoelastic properties differently than those observed in non-human species [59,60]. This gap in human studies is a critical issue, as the results from animal models may not always accurately represent the clinical challenges encountered in human ophthalmology, particularly when it comes to understanding diseases such as anomalous PVD, macular holes, and other vitreous- related retinal conditions.

Most of the studies included in this review used ex vivo methods, where the vitreous body was removed from the eye and tested outside of its natural biological environment. While ex vivo studies can provide valuable insights into the basic rheological properties of vitreous humor, they do not account for the complex interactions that occur in vivo, including the effects of ocular pressure, the mechanical forces during eye movement, and the interplay between the vitreous and other ocular structures [7,61]. The dearth of in vivo human studies limits our understanding of how vitreous viscosity may change under physiological and pathological conditions, and how these changes may impact clinical outcomes.

### Clinical Implications

As outlined above, the heterogeneity in experimental models, rheometric devices, and outcome measures limits the immediate clinical use of current viscosity data. Quantifying vitreous viscosity preoperatively or intraoperatively could allow surgeons to tailor management of vitreous-related pathologies, but without standardized, human-specific values, these measurements cannot yet reliably guide surgical planning

For example, in vitreomacular traction, inconsistencies in viscosity measurement methods and lack of in vivo human data make it difficult to predict whether the vitreous will remain stable or progress to detachment. More viscous vitreous may require different handling than less viscous vitreous, influencing instrument choice and surgical approach. In retinal detachment, the absence of validated human viscosity ranges means decisions about whether a scleral buckle alone is adequate, or if adjunctive vitrectomy is required, remain based on indirect evidence [36].

These same limitations affect intravitreal drug delivery models. Reported steady-state viscosity values span several orders of magnitude depending on technique and sample preparation, which could lead to errors in estimating drug dispersion or clearance rates. Standardized, in vivo human viscosity measurements would enable optimization of drug formulation, injection volume, and delivery site for improved pharmacokinetics and therapeutic effect [62].

In cases of vitreous loss due to trauma, surgery, or disease, the development of an ideal vitreous substitute that mimics the viscosity and viscoelastic properties of natural vitreous is a significant challenge [9,13,63]. Current designs are often benchmarked against ex vivo data that may not accurately reflect human in vivo properties. Without robust human reference values, substitutes risk mechanical mismatches that could compromise long-term retinal support or stability. Minimally invasive human studies are essential before viscosity metrics can reliably guide substitute development.

### Limitations

This systematic review has several limitations that merit consideration. First, the quality and heterogeneity of the included studies present challenges for synthesis. Many studies used differing rheometric devices, experimental models, and viscosity metrics, which precluded a quantitative meta-analysis and limited direct comparability. The scarcity of human studies further constrains the generalizability of the findings, as the majority of included studies used animal models with different vitreous composition and aging characteristics. Moreover, much of the data was derived from ex vivo experimentation, which does not fully account for the biomechanical dynamics of the in vivo environment.

Limitations in the review process also stem from language and publication bias. Only peer-reviewed, English-language studies were included, potentially excluding relevant data published in other languages or gray literature. Although dual-reviewer screening and data abstraction minimized the risk of selection bias, the lack of available data in some studies—particularly those omitting key experimental variables or failing to report viscosity outcomes in standardized units—introduces variability and limits reproducibility. Finally, while risk of bias tools were used to evaluate study quality, many studies lacked sufficient reporting detail, resulting in several domains being rated as “unclear.” These limitations highlight the need for future research to adopt standardized reporting frameworks and to prioritize in vivo human investigations.

### Technological Advancements and Future Directions

The review of existing studies reveals a pressing need for the development of more precise, accurate, and minimally invasive techniques to measure vitreous viscosity in vivo, particularly in human eyes.

Although a variety of rheometric devices, such as capillary tube viscometers, rotational viscometers, and cone-plate geometry, have been employed, these methods have limitations in terms of invasiveness, accuracy, and applicability to human clinical settings. For instance, many of the devices used in the studies reviewed are not feasible for routine clinical use, and their ability to provide real-time measurements of vitreous viscosity is limited.

The future of vitreous rheometry lies in the development of advanced, portable devices capable of measuring vitreous viscosity in vivo with minimal disruption to the ocular environment. Such devices would need to be non-invasive or minimally invasive to ensure patient comfort and safety, while also providing accurate and reproducible results. One potential avenue for development is the use of optical coherence tomography (OCT)-based techniques or ultrasound elastography, both of which have been successfully used in other areas of ophthalmology to assess tissue properties [64–69]. These technologies could be adapted for use in measuring vitreous viscosity, offering a non-invasive means of obtaining real-time measurements of vitreous properties.

In addition to technological advancements, future research should focus on addressing the lack of data regarding the physiological and pathological changes in vitreous viscosity. Studies should aim to investigate how vitreous viscosity changes with age, disease, and therapeutic interventions. For example, in patients with anomalous PVD or other vitreoretinal interface diseases, understanding how the viscosity of the vitreous changes over time could help identify early biomarkers of disease progression and improve the timing of interventions. Similarly, exploring how vitreous viscosity is affected by conditions such as diabetes or glaucoma could lead to better-targeted treatments for these conditions [70–72].

Finally, future research should aim to develop standardized protocols for measuring vitreous viscosity, as the variability in study methodologies, mathematical models used, and experimental assumptions identified in this review makes it difficult to compare results across studies. A consensus on the optimal rheometric techniques and device types for measuring vitreous viscosity would help standardize measurements and improve the reliability of research findings. Additionally, multi-center studies focusing on human eyes are urgently needed to validate current findings and expand our understanding of vitreous rheology in health and disease.

## Conclusion

This systematic review highlights the critical need for more research into the viscosity of human vitreous humor. While significant progress has been made in studying the viscoelastic properties of vitreous humor in animal models, there remains a lack of in vivo human studies and standardized methods for measuring vitreous viscosity. The development of advanced, minimally invasive rheometric techniques is essential to advancing our understanding of vitreous physiology and its clinical implications. With better tools to quantify vitreous viscosity, clinicians could optimize the management of vitreous-related pathologies, enhance intravitreal drug delivery, and improve the design of vitreous substitutes. Ultimately, addressing the gaps in our understanding of vitreous viscosity could lead to more effective treatments and improved patient outcomes in ophthalmology.

## Data Availability

All data produced in the present work are contained in the manuscript.

## Acknowledgments

None

## Commercial Relationships

None

## Author Contributions

Lauren Ong: [Conceptualization, Data Curation, Formal Analysis, Investigation, Methodology, Resources, Validation, Visualization, Writing – Original Draft Preparation, Writing – Review & Editing]; Tara Suresh: [Data Curation, Investigation, Validation, Writing – Original Draft Preparation, Writing – Review & Editing]; Esther Son: [Data Curation, Investigation, Validation, Writing – Original Draft Preparation, Writing – Review & Editing]; Daniel Schwartz: [Conceptualization, Investigation, Validation, Supervision, Writing – Review & Editing]; Frank Brodie: [Conceptualization, Investigation, Validation, Supervision, Writing – Review & Editing]

## Supporting information captions

**S1 Appendix.** Detailed Search Strategy. A comprehensive breakdown of the search terms, Boolean operators, and database-specific queries used for the systematic review.

**S2 Appendix.** PRISMA 2020 Checklist.

**S3 Appendix.** Risk of Bias Assessment. A summary of the quality assessment results for included studies, utilizing SYRCLE’s risk of bias tool for controlled studies and an adapted Newcastle-Ottawa Scale (NOS) for animal studies.

**S4 Appendix.** Summary of Included Studies. A detailed table outlining the key characteristics of the included studies.

