## Appendix S1 for "Insight into vitreous biomechanics and clinical implications: A systematic review"

**PubMed (11/23/2023) - 1,452 results**

("Vitreous Body"[MeSH] OR "Retina"[MeSH]) AND ("Viscosity"[MeSH] OR "Rheology"[MeSH] OR "Biomechanical Phenomena"[MeSH] OR "shear" OR "oscillation")

**Embase (11/23/2023) - 498 results**

(‘vitreous body’) AND (‘viscosity’ OR ‘rheology’ OR (‘biomechanical phenomena’) OR ‘shear’ OR ‘oscillation’)

**Scopus (11/23/2023) - 451 results**

( vitreous AND body ) AND ( viscosity OR rheology OR ( biomechanical AND phenomena ) OR shear OR oscillation )
