## Appendix S2 for "Insight into vitreous biomechanics and clinical implications: A systematic review"

| **Domain** | **Selection** | **Ascertainment: Exposure** | **Ascertainment: Outcomes** | **Causality: Alternatives** | **Causality: Challenge/Rechallenge** | **Causality: Dose-Response Relationship** | **Causality: Follow-Up Period** | **Reporting: Study Details** |
| --- | --- | --- | --- | --- | --- | --- | --- | --- |
| Abdelkawi SA et al 2012 | Yes | Yes | Yes | Unclear | No | N/A | Yes | Yes |
| Abdelkawi, S.A. 2014 | Yes | Yes | Yes | Unclear | No | N/A | Yes | Yes |
| Aguayo J et al, 1985 | Yes | Yes | Yes | Unclear | No | N/A | Yes | Unclear |
| Balazs, E.A. et al, 1959 | Yes | Yes | Yes | Unclear | No | N/A | Unclear | No |
| Berman, E.R. 1963 | Yes | Yes | Yes | Unclear | No | N/A | Yes | Yes |
| Berman, E.R. and Michaelson, I.C. 1964 | Yes | Yes | Yes | Unclear | No | N/A | Yes | Yes |
| Bettelheim FA; Wang TJ 1976 | Yes | Yes | Yes | Unclear | No | Yes | Yes | No |
| Boruchoff, S.A.; Woodin, A.M. 1956 | Unclear | Yes | Yes | Unclear | No | N/A | Yes | No |
| Chakrabarti B; Hultsch E 1976 | Unclear | Yes | Yes | Unclear | No | Yes | Yes | No |
| Colter J et al 2015 | Yes | Yes | Yes | Unclear | No | Yes | Yes | Yes |
| Connelly K et al 2016 | Unclear | Yes | Yes | Unclear | No | Yes | Yes | Unclear |
| Elmali, A. et al 2021 | Yes | Yes | Yes | Yes | No | Yes | Yes | Yes |
| Filas BA et al 2014 | Yes | Yes | Yes | Yes | No | Yes | Yes | No |
| Gisladottir S et al 2009 | Unclear | Yes | Yes | Yes | No | Yes | Yes | No |
| Hajjarian Z; Nadkarni SK 2013 | Unclear | Yes | Yes | Yes | No | Yes | Yes | Unclear |
| Huang D et al 2018 | Yes | Yes | Yes | Yes | No | Yes | Yes | Yes |
| KAWANO, S.‐I. et al 1982 | Yes | Yes | Yes | Yes | No | Yes | Yes | Yes |
| LARSEN G 1958 | Yes | Yes | Yes | Yes | No | Yes | Yes | Yes |
| Lerman S et al, 1984 | Yes | Yes | Yes | Yes | No | Yes | Yes | Yes |
| Loch, C. et al 2012 | Yes | Unclear | Unclear | Yes | No | Unclear | Yes | No |
| Locke JC and Morton WR 1965 | Yes | Yes | Yes | Yes | No | Yes | Yes | Yes |
| Mensitieri, M.; et al 1994 | Yes | Yes | Yes | Unclear | No | Yes | Yes | Yes |
| Neal RE et al 2005 | Yes | Yes | Yes | Yes | No | Yes | Yes | Yes |
| Nickerson CS et al 2008 | Unclear | Yes | Yes | No | No | Yes | Yes | Unclear |
| Piccirelli, M. et al 2012 | Yes | Yes | Yes | Unclear | No | Yes | Yes | Yes |
| Pokki J et al 2012 | Unclear | Yes | Yes | Yes | No | Yes | Yes | Unclear |
| Pokki J et al 2015 | Unclear | Yes | Yes | Yes | No | Yes | Yes | Yes |
| Rangchian A et al 2019 | Yes | Yes | Yes | Yes | No | Yes | Yes | Yes |
| Rangchian, A. et al 2020 | Yes | Yes | Yes | Yes | No | Yes | Yes | Yes |
| Schulz, A. et al 2019 | Yes | Yes | Yes | Yes | No | Yes | Yes | Yes |
| Shafaie S et al 2018 | Yes | Yes | Yes | Yes | No | Yes | Yes | Yes |
| Sharif-Kashani P et al 2011 | Yes | Yes | Yes | Unclear | No | Yes | Yes | Unclear |
| Srikantha N et al 2022 | Yes | Yes | Yes | Unclear | No | Yes | Yes | Yes |
| Suri, S and Banerjee, R 2006 | Unclear | Yes | Yes | Yes | No | Yes | Yes | Unclear |
| Swindle KE et al 2008 | Unclear | Yes | Yes | Unclear | No | Yes | Yes | No |
| Thakur SS et al 2020 | Unclear | Yes | Yes | Unclear | No | Yes | Yes | Unclear |
| Tokita M et al 1984 | Unclear | Unclear | Unclear | Unclear | Unclear | Yes | Yes | Unclear |
| Wang Z et al 2013 | Unclear | Yes | Yes | Yes | No | Yes | Yes | Unclear |
| Weber H et al 1982 | Yes | Yes | Yes | Unclear | No | Yes | Yes | Yes |
| WOODIN AM; BORUCHOFF SA 1955 | Yes | Yes | Yes | Unclear | No | Yes | Yes | Yes |
| Yoon S et al S 2013 | Unclear | Yes | Yes | Yes | No | Yes | Yes | Yes |
| Zhang, Q et al, 2014 | Yes | Yes | Yes | Yes | No | Yes | Yes | Yes |
