## Appendix S3 for "Insight into vitreous biomechanics and clinical implications: A systematic review"

| **Study Name** | **Patient/Experimental Demographics + Conditions** | **Viscosity Measurement Device Name** | **Device Type** | **Device’s Viscosity Controlled Variable + Test Range(s)** | **Other Special Conditions for Viscosity Testing** | **Viscosity Outcome Variable (value/range + units)** | **Mathematical Equation/Model for Viscosity** | **Reported Viscosity Complications/Limitations/Notes** |
| --- | --- | --- | --- | --- | --- | --- | --- | --- |
| Abdelkawi SA et al 2012 | 21 New Zealand albino male rabbits weighing 2-2.5kg | “Brookfield DV-III, Engineering Laboratory, USA”  viscometer | Cone-plate | Number of cone rotations per minute from 1-50 rpm (thus shear rates of 7.5-375 s^-1) | N/A | Viscosity range:  5.2 × 10−3 − 17.5 × 10−3 Pa.s at  a shear rate of 7.5 s−1    to  0.96 × 10−3 − 1.92 × 10−3 Pa.s at a shear rate of 375 s−1 | F = mS^n  F = shear rate  S = shear stress  m = consistency  n = fluid flow index | N/A |
| Abdelkawi, S.A. 2014 | 39 New Zealand male rabbits weighing 2-2.5kg (3 control) | Digital viscometer type ‘‘Brookfield DV-III, Eng. Lab.,  USA’’ | Cone-plate | Shear rate varying from 7.5 to 374 s^-1 | N/A | Viscosity range:  m 5.2 to  17.5 cp at a shear rate of 7.5 s-1  to  0.96–1.92 cp at a shear rate of 375 s-1 | F = mS^n  F = shear rate  S = shear stress  m = consistency  n = fluid flow index | N/A |
| Aguayo J et al, 1985 | Bovine vitreous | Wells Brookfield Model LVT 17961  cone-plate viscometer (Brookfield Engineering Laboratories, Inc; Stoughton, MA) | Cone-plate | N/A | Temperature held constant at 22*C | Viscosity after 12 hr at 22*C  Shear rates  230: 6.4 ± 1.0  115: 7.2 ±2.1  46: 9.5 ± 5.0  23: 15.7 ± 5.3  11.5: 21.1 ±9.9 | Viscosity is a function of shear rate and shear  stress | The revolutions  per minute of the cone reflects the shear rate  “Because the flow and viscosity data in  this report describe non-Newtonian fluids, "apparent"  viscosity is actually being measured.” |
| Balazs, E.A. et al, 1959 | Owls | Ostwald capillary viscometer | Capillary tube | High-speed centrifugation (105,000 x *g*) | 37 degrees | Specific viscosity: 8.20  After 1hr centrifugation: 6.86 | Specific viscosity: 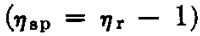 | N/A |
| Berman, E.R. 1963 | 28 cattle eyes 30 min after death, frozen at -15*C | Cannon-Manning semimicro capillary viscometers at  30°C | Capillary tube | No. 150 viscometer (flow time for water, 20.8 sec)  Very high viscosity samples: No. 200 viscometer (flow  time for water, 7.5 sec). | Samples dialysed overnight against 0.2M NaCl (without buffer)  Experiments at room temperature (10-18*C) | Cortical preparations (HA isolated): 420-1800 (η in ml/g)  Central portion (HA isolated): 510-3320 (η in ml/g)  The corresponding, values of the original cortical and central  preparations were 820 and 1360, respectively | N/A | “No pretreatment of the preparations, other than centrifugation and dialysis, was required, and elution from the column was accomplished at  neutral pH.” |
| Berman, E.R. and Michaelson, I.C. 1964 | 24 human post-mortem eyes | Cannon-Manning semimicro capillary viscometer | Capillary tube | Not mentioned | Measured at 30*C | Relative viscosity (cortical)  Age 0-2 years: 1.04  13-45 years: 1.53±0.19  50-85 years: 2.08±0.28  Myopic eyes: 1.54±0.10  Relative viscosity (central)  Age 0-2 years: 1.03  13-45 years: 1.59±0.21  50-85 years: 1.83±0.12  Myopic eyes: 1.44±0.15  Viscosity numbers of the central  portion were consistently higher than those of the cortical layer | Viscosity numbers were calculated from the concentrations of hyaluronic acid (in g/ml) | N/A |
| Bettelheim FA; Wang TJ 1976 | Adult bovine eyes and calf eyes, <1 day postmortem, kept at 4*C | “Vibron” direct-reading dynamic viscoelastometer model DDV-II (Toyo Measuring  Instrument Co. Ltd, Tokyo, Japan) | Viscoelastometer | N/A | 2 compression chucks inserted into vitreous  “Instead of clamping the specimen, we used U-shaped aluminum chucks as a compression cage. The whole eyeball was encased in a mold made of approximately 1 cm-thick “Kerr Permlastic:” used by dentists to make impressions for dentures, etc” | Unclear | Unclear | N/A |
| Boruchoff, S.A.; Woodin, A.M. 1956 | In vivo: adult rabbits under nembutal anesthesia  Pooled rabbit’s vitreous: stored in frozen state | In vivo: horizontal viscosimeter  Pooled rabbit vitreous: Couette viscosimeter | In vivo: horizontal viscosimeter  Rotational | Constant pressure apparatus, shear rates of 35-500 sec-1 | Measurements made at 30*C | Unclear | Shear rate:  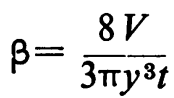  “Relative viscosity was obtained by comparing  the flow time of vitreous with that of saline at the same shear rate” | N/A |
| Chakrabarti B; Hultsch E 1976 | Liquid vitreous from anesthetized owl monkeys | Cannon-Ubbelohde semi-microdilution viscometer | Capillary tube | Molecular weight | “All optical and chemical measurements were performed after extensive dialysis of the vitreous samples against 0.15 M NaCl at 4*C.” | Intrinsic Viscosity [η]  Sample I: 4450 cc/gm  Sample II: 4800 cc/gm  Sample III: 4300 cc/gm  Sample IV: 220 cc/gm  Sample V: 4200 cc/gm | Molecular weight (M) calculated from plot of intrinsic viscosity (η) against known molecular weight samples on the basis of Mark-Houwink relationship, [η] = KM^a | N/A |
| Colter J et al 2015 | Premature (n=13), infant (n=11), and adult (n=6) sheep collected post-mortem, stored in phosphate buffered saline (PBS) | AR-G2 rheometer (TA Instruments, New Castle, DE) with accompanying Rheology Advantage Instrument Control software | Rotational | N/A | “The linear viscoelastic region (LVR) was established for each material by subjecting preliminary samples (n¼3) to a strain sweep over three decades (0.1–100%) at 1 Hz” | Unclear | N/A | N/A |
| Connelly K et al 2016 | Porcine (n=25) and bovine (n=40) eyes, tested within 8 hours of delivery | TA instruments AR 2000 controlled stress rheometer with probe geometry attached. | Probe | Creep tests performed for 5 minutes with applied shear stress of 30 Pa | All testing at room temperature, 20*C | Steady state viscosity and compliance intercept average values for porcine and bovine vitreous samples calculated from creep curves:  Bovine ηss [Pa-s]: 1.43 × 10^5 ± 2.19 × 10^4  Porcine ηss [Pa-s]: 4.35 × 10^4 ± 4.80 × 10^3 | 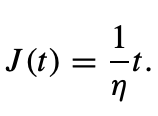  J = compliance  t = time  J(t) = creep curve (time dependent deformation of the sample) | “Creep curves generated by probe measurements in an infinite Newtonian fluid may be used to calculate viscosity at steady state.” |
| Elmali, A. et al 2021 | 12 male New Zealand white rabbits, 6-8 months old | Brookfield DV2T RV viscometer and CPA-40 Z cone | Rotational and cone-plate attachment | 1, 5, 10, 20, 50, 100 rpm results were  evaluated | All testing at 25*C | Control samples:  10 rpm median viscosity: 4.74 cP (range; 1.96–33.35 cP)  20 rpm median viscosity: 3.68 cP  (range; 0.65–8.18 cP) | N/A | N/A |
| Filas BA et al 2014 | Fresh bovine and porcine eyes collected immediately after animal sacrifice | Cleated stage of a stress-controlled AR-G2 rheometer (TA Laboratories, New Castle, DE; Fig. 1C) | Rotational | N/A | N/A | Unclear | Oscillatory sweeps applied a frequency-dependent, sinusoidal stress: 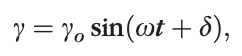  Measure of elastic (G’) relative to viscous (G”) behavior or material:  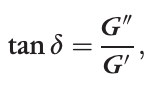 | N/A |
| Gisladottir S et al 2009 | Porcine eyes, dissected vitreous humour | Brookfield digital DV-I+rotational viscometer (Brookfield Engineering Laboratories, Inc., Middleboro,  MA, USA) | Rotational | Gel vitreous measured with 0.3, 0.6 and 1.5 rpm  Liquid vitreous at 6, 12 and 30 rpm | N/A | Liquid vitreous viscosity: 6.29±2.3 cp  Liquid vitreous humour is about six times more viscous than saline | Stokes-Einstein equation: diffusion inversely correlated with viscosity of the medium | N/A |
| Hajjarian Z; Nadkarni SK 2013 | Frozen bovine synovial fluid and vitreous humor | Laser Speckle Rheology (LSR)  Light from He-Ne Laser  (633 nm) was coupled into a single mode fiber (SM600)  High frame rate CMOS camera (PL761, Pixelink, Ontario, Canada)  Reference standard mechanical rheometer (ARG2, TA Instruments, MA) | Laser speckle rheology  Rotational | Reference standard rheometer: shear oscillatory torque (stress) over frequency range 0.1-100 Hz | N/A | \|G*(ω)\| curves (Figure 9) | Mean square displacement of light scattering centers (MSD)  Generalized Stokes’-Einstein Relation (GSER) deduces viscoelastic modulus, G*(ω) from measured MSD | N/A |
| Huang D et al 2018 | Fresh bovine eyes transported on ice | Stress-controlled frequency sweeps:  oscillation  rheometer (Physica model UDS-200, Parr Physica Inc., Stuttgart,  Germany) with a 25 mm-radius plate | Oscillation | Oscillatory frequency sweeps (0.63–6.28 rad/s)  at a fixed shear strain amplitude (γ0 = 3%) were performed to measure the dynamic shear modulus | N/A | Measured: storage modulus (G’) and loss tangent (tan δ)  Bovine vitreous storage modulus: 7.37 ± 5.04 | Measure of elastic relative to viscous behavior of material and shows damping capacity: 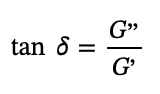 | N/A |
| KAWANO, S.‐I. et al 1982 | Human vitreous from post mortem eyes enucleated for keratoplasty  (9 eyes of 6 patients)  10 pig eyes | Cannon-Fenske kinematic and cone-plate rotary  viscometers | Capillary tube  Cone-plate | N/A | Thimerosal (methiolate sodium) in a concentration of 0.005% was added to the vitreous for long term observations. It was confirmed that the addition of this amount of thimerosal did not affect the viscosity of the excised vitreous. | Unclear | Kinematic Viscosity (Stokes) X Density = Absolute Viscosity (Poise) | N/A |
| LARSEN G 1958 | 244 rabbits | Special  viscosimeter of Ostwald's type, consisting  of a capillary tube 0.29 mm. in diameter | Capillary tube | Viscosimeter was kept in a thermostat at a constant temperature of 30C | One drop of 0.5% diethyldithiocarbamate and toluene was added to prevent a possible depolymerization by copper ions and ascorbic acid or any other decomposition | Relative Viscosity  Left eyes: range 1.113-1.196; 1.144±0.002  Right eyes: range 1.100-1.204; 1.145±0.003 | N/A | N/A |
| Lerman S et al, 1984 | 4 hybrid monkey, 26 rabbit eyes | Modified viscometer | Modified viscometer | Viscometry analyses at 35*C | N/A | Viscometry (secs)  Rabbits weighing <2.5kg (R.E./L.E.): 415/372, 511/563, 458/420, 510/532, 383/408 | N/A | N/A |
| Loch, C. et al 2012 | Human and porcine vitreous humor | Rotation viscosimeter  (RVDV-II+CP, Brookfield Engineering Laboratories,  Massachusetts, USA) | Rotational with cone-plate geometry | N/A | N/A | Human/porcine viscosity: 606.0 ± 9.6 | Not mentioned | N/A |
| Locke JC and Morton WR 1965 | 176 samples from 97 donors (human) | Ostwald type viscosimeter pipette with a 0.5-mm. diameter | Capillary tube | Flow time | 37*C | Average: 1.97  Median: 1.86  Range: 10.6 to 5.50 | Relative viscosities were measured, with distilled water the reference fluid | N/A |
| Mensitieri, M.; et al 1994 | Goat (caprine), sheep (ovine), pig (suine), calf (bovine), rabbit vitreous bodies | Bohlin VOR Rheometer  (Bohlin Reologi AB, Lund, Sweden) | Rotational | Strain sweep tests at a fixed oscillation frequency (consisting in monitoring the viscoelastic properties while logarithmically varying the strain amplitude γ0) 0.05-10 Hz | T = 37*C | The strain sweep tests at fixed frequency show a  linear behaviour for G' and G" versus γ0 up to about  0.01 strain | 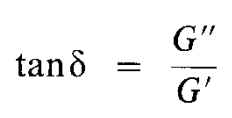 | N/A |
| Neal RE et al 2005 | 2 pairs of phakic/pseudophakic donor globes, 1 pseudophakic glove, 5 phakic donor eyes, frozen on dry ice within 4-7 hr post-mortem | Cannon-Ubbelhode Calibrated Semi-Micro Viscometer | Capillary tube | Efflux times were obtained with three  different dilutions (between 2.5 and 10-fold dilution) for  each sample | N/A | 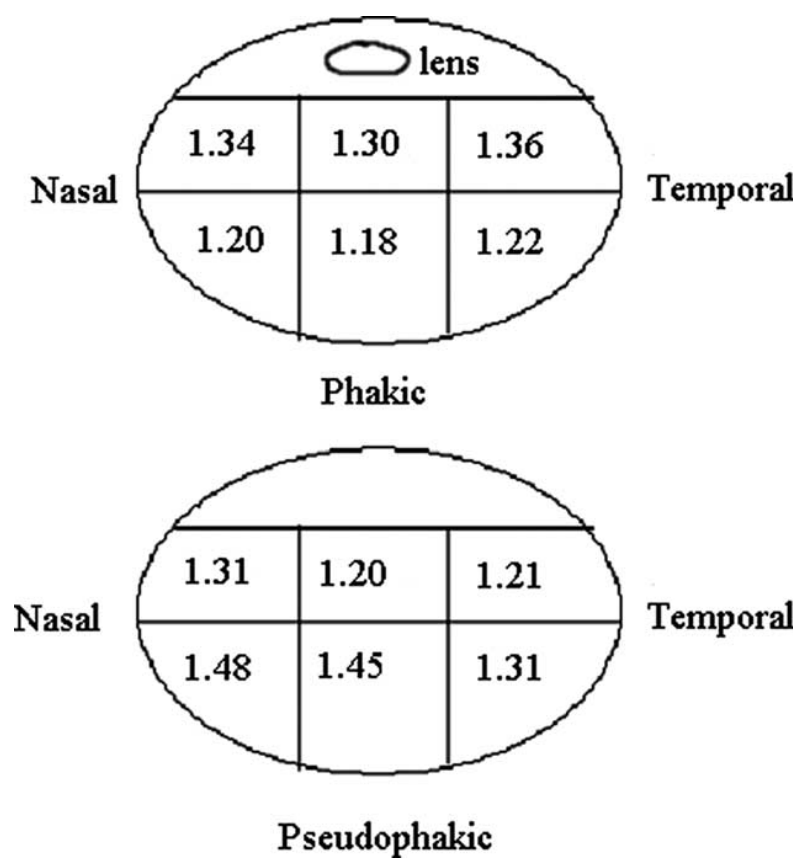 | Viscosity coefficients are dependent upon the concentration of the macromolecules present | N/A |
| Nickerson CS et al 2008 | Fresh bovine and porcine eyes tested between 3 and 48 h postmortem | Oscillatory: ARES-RFS fluids  rheometer from TA Instruments, Inc. (New Castle, DE, USA) using the  ‘‘cleat geometry’’ to overcome slip  Steady shear rate tests | Cleat geometry | Oscillatory: Oscillatory strain  (γ = 3%) at a fixed frequency (ω = 10 rad/s) for up to 90 min  Shear: Steady shear rate (γ = 0.1s^-1) applied for 150s while continuously monitoring the resulting shear stress | Closed, humid atmosphere at 20*C and with zero normal force on the samples | Average steady-state moduli for bovine vitreous: G’fin = 7.0±2.0 Pa and G’’fin = 2.2±0.6 Pa  Porcine vitreous: G’fin = 2.8±0.9 Pa and G’’ fin = 0.70±0.4 Pa | n/A | N/A |
| Piccirelli, M. et al 2012 | Right eyes of 19 healthy subjects | N/A | Magnetic resonance imaging | N/A | N/A | Unclear | N/A | N/A |
| Pokki J et al 2012 | Porcine vitreous humour | AFM colloidal cantilever  Magnetic microrobot | Colloidal cantilever  Magnetic robot | Varied constant velocities; force response recorded  Stepwise constant  force (±2.3 µN and ±4.6 µN) | N/A | AFM:  k (elasticity): 8.1 ×10−1 (±4.7 × 10−1 ) [Pa m]  γ0 (steady state velocity): 1.3x10^-2 (±0.9 × 10−2) [Pa s m] | 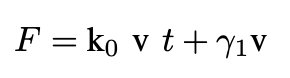 | N/A |
| Pokki J et al 2015 | Human donor eyes  Anesthetized young New Zealand albino rabbits | Magnetically actuating a microprobe and tracking its position | Magnetic microprobe | Varied constant velocities; force response recorded  Stepwise constant  force (±2.3 µN and ±4.6 µN) | N/A | Ex vivo human vitreous (mean±SD values: γ0 = 3.6±0.9 Pa·s; k1 = 1.6±0.3 Pa)  Porcine vitreous (mean±SD values: γ0 = 40.4±14.5 Pa·s; k1 = 42.6±39.3 Pa) | 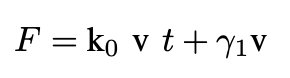 | N/A |
| Rangchian A et al 2019 | 38 porcine eyes | Stress-controlled shear rheometer (TA instruments, AR-2000) using patented probe | "Patented probe" | Creep test: Constant torque is applied | Eye was placed on a 3D printed cube to hold the eye in a secure position to avoid any movement during the experiment | Angular displacement of flow is recorded  Group 2 (control group) 15.4×10^3  [Pa.s] | 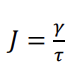 | N/A |
| Rangchian, A. et al 2020 | 44+18 fresh porcine eyes | Stress-controlled  rheometer (TA instruments, AR 2000) with a 0.87 mm  diameter cylindrical probe | Cylindrical probe | Torque of 1 lNm with zero normal force over 6 min | N/A | Viscosity at steady state: slope of the linear segment of creep curve between t = 240 s and t = 300 s  Group 1 pre-injection: 6 6x10^-5 [1/Pa.s]  Group 2 all time intervals: 4x10^-5 to 5x10^-5 [1/Pa.s] | Creep compliance (J)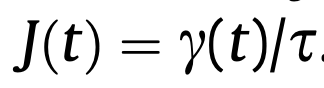 | N/A |
| Schulz, A. et al 2019 | 190 human eyes  Bovine and porcine eyes | Rotational rheometer Physica MCR 101  (Anton Paar, Graz, Austria) with a parallel plate  geometry and a Peltier element | Rotational with parallel plate geometry | Frequency  sweep between 0.1 and 100 s^-1 | N/A | Younger donors: G’ = 1.4 to 14.5 Pa (log(G’) = 0.14-1.16 Pa) and G’’ = 0.55  to 7.39 Pa (log(G’’) =-0.26–0.87 Pa)  Older donors: G’ = 0.24 to 6.60 Pa  (log(G’) = 0.62–0.82 Pa) and G’’ = 0.12 to 4.04 Pa  (log(G’’) = -0.91–0.61 Pa) | Rheological properties are determined as storage modulus  (G’ ) and loss modulus (G’’) via dynamic mechanical  analyses/frequency sweep tests | 500 grit sandpaper  was used to reduce the wall slip |
| Shafaie S et al 2018 | Bovine, porcine, and ovine eyes  10 human eyes | Oscillatory stress sweep (OSS): AR-G2 AR 1500ex rheometer (TA instruments) with 40 mm parallel plate and a geometry  gap of 600 mm  Frequency stress sweep (FSS) | Parallel plate | Oscillation from 0.01 to 100 Pa at 1 Hz  Frequency sweep from linear viscoelastic region (LVR) of stress sweep between 0.1 and 10 Hz | N/A | OSS:  Human: 1.4 ± 0.95 Pa  Bovine: 1.7 ± 0.31 Pa  Porcine: 1.4 ± 0.14 Pa  FSS:  Ovine: G′ (6.7 ± 0.48 Pa), G″ (3.7 ± 0.45 Pa), complex viscosity (12.3 ± 0.94 Pa.s) | Storage (G′) and loss (G″) moduli and the magnitude of the complex viscosity  [ƞ*] | N/A |
| Sharif-Kashani P et al 2011 | Porcine eyes | Stressed-controlled shear rheometer (AR-2000, TA Instruments) with 20 mm  parallel disc geometry | Parallel plate | Creep compliance: concentrations 0.5, 1, 3, 5 mg/ml for constant shear stresses of τ0=0.5, 1, and 2 Pa | N/A | Averages for the storage and loss modulus are  G’=1.08±0.22 Pa and G’’=0.25±0.07  Average storage modulus from Voigt-Kelvin element: G’1=1.66±0.96 Pa  G’2=1.14±0.71 Pa | Creep compliance J(t)  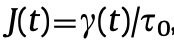  Viscoelastic spectra model (Voigt-Kelvin) to model creep behavior  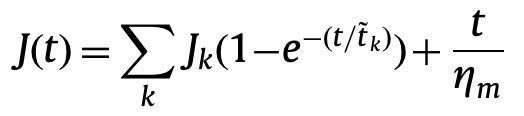 | N/A |
| Srikantha N et al 2022 | Porcine eyes | Fluorescence Recovery After Photobleaching (FRAP)-labeled molecules | Fluorescence Recovery After Photobleaching | FITC-labelled molecules (λem = 510 nm) | N/A | Unclear | Diffusion coefficient 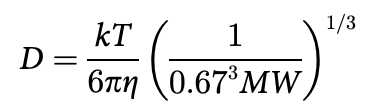 | N/A |
| Suri, S and Banerjee, R 2006 | Goat eye | Concentric cylinder  viscometer (Contraves low shear 30 viscometer,  Zurich) | Concentric cylinder | Shear rate | N/A | “Very high”  Greater than 4000 cP at a shear rate of 0.15 sec^-1 | Viscosity is  measured as a function of shear rate | N/A |
| Swindle KE et al 2008 | Porcine eyes from 6-month-old pigs | Vilastic-3 oscillatory capillary tube rheometer (Vilastic, Austin,  TX) | Capillary tube | 25*C at 2 Hz, with shear rate increasing from 0.1 to 100 s^-1 | N/A | At frequency 2 Hz and shear rate 0.1s^-1, storage modulus: 3.46 ± 0.30 Pa, loss modulus: 0.71 ± 0.12 Pa  After shear thinning, final values:  Storage: 0.64 ± 0.18 Pa  Loss moduli: 0.37 ± 0.06 Pa | Elastic moduli (E)  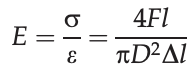 | N/A |
| Thakur SS et al 2020 | Porcine eyes (n = 6) | Discovery HR-2  rheometer (TA Instruments, New Castle, DE, USA) equipped with a  40 mm parallel plate geometry | Parallel plate | During frequency sweeps, gels were held at a constant strain of 3%  and subjected to an angular frequency (ω) of 10 rad/s | N/A | standard error of the estimate  for G’ = 0.21 Pa, G” = 0.10 Pa, η* = 0.02 Pa s | 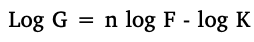  G = shear rate  F = shear stress  n = pseudoplasticity index  K = flow consistency index | N/A |
| Tokita M et al 1984 | Bovine eyes | Torsion pendulum  apparatus | Torsion pendulum  apparatus | Frequency of 0.1 to 0.003 Hz | N/A | Shear modulus: order  5dyn/cm^2 at 20*C | 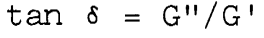 | N/A |
| Wang Z et al 2013 | Porcine vitreous humor | A 0.55mm-diameter NdFeB sphere [Supermagnete, Germany] was driven by magnetic fields to move inside the vitreous humor | Magnet | Cycled stepwise constant magnetic force (±4.6µN) | N/A | Graph | 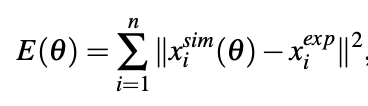 | N/A |
| Weber H et al 1982 | 100 pig eyes | Computer analysis of  the resonant curves | Computer analysis of  the resonant curves | N/A | Pig eyes put into physiological NaCl solution after enucleation | Unclear | N/A | N/A |
| WOODIN AM; BORUCHOFF SA 1955 | Ox vitreous humor | Couette viscosimeter  The inner cylinder, diameter 12 mm., was immersed to a depth of 10 cm. in 10 ml. of solution | Rotational | Shear rate varied from 0.5 to 150 sec^-1 | N/A | Fresh gel homogenate  Nrel at β = 30 sec^-1: 1.82 - 2.0  Flow behavior at low shear rate: Anomalous (versus Newtonian)  Hydroxyproline content (ug/ml): 6.0 - 8.75 | “Specific viscosity”: 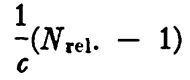  “Intrinsic viscosity”: 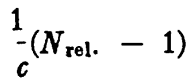  “Reduced viscosity: (Nrel - 1) | N/A |
| Yoon S et al S 2013 | Porcine eye globes | Size of the microbubble was monitored by an optical microscope (AM411T,  Dino-Lite, Torrance, CA) operating at 230× magnification | Microbubble | N/A | N/A | Young’s moduli anterior: 34.0 ± 1.3 Pa  Shear viscosity: 0.052 ± 0.003 Pa·s  Young’s moduli central: 8.3 ± 0.2 Pa  Shear viscosity: 0.035 ± 0.005 Pa·s | Young’s modulus (*E*) and shear viscosity (η) were found by measuring tmax and the decaying profiles of the microbubble | N/A |
| Zhang, Q et al, 2014 | Bovine (in situ and in vitro) | AR G2 rheometer (TA Laboratories, New Castle, DE, USA) | Parallel plate | Stress-controlled frequency sweep (ω = pi/5 to 2pi rads/sec) at fixed shear strain amplitude (γ0 = 3%) | In situ: trypsin, mimics, or mixture of mimics plus trypsin injected  In vitro: collegenase and mimics  A cleated, parallel plate (20-mm diameter) was used to prevent slippage | Controls: G’ = 11.1 ± 2.9 Pa | Storage modulus (G’, Pa), parameter for evaluating stiffness of the vitreous gel | N/A |
